# Real-Time EEG-Based Epileptic Seizure Prediction Using Artificial Intelligence: A Systematic Review

**DOI:** 10.1101/2025.10.09.25337692

**Authors:** Zikang Song, Kim Arrowsmith, Dion Henare, Mangor Pedersen

## Abstract

**Background:** Epilepsy affects approximately 50 million people worldwide, and seizures remain difficult to predict in onset, severity, and duration. Real-time seizure prediction may enable proactive intervention and improve patient safety and quality of life. Despite the development of high-performing algorithms, translation remains limited by predictive accuracy, interpretability, and generalisability.

**Objectives:** This systematic review evaluates artificial intelligence (AI) models for real-time epileptic seizure prediction and assesses machine learning and deep learning performance alongside real-time responsiveness, interpretability, and multimodal integration.

**Methods:** We searched PubMed, Scopus, IEEE Xplore, and ScienceDirect (1 Jan 2017–31 May 2025); 23 studies met eligibility. Inclusion required EEG-based real-time prediction with model/dataset/validation details and sufficient methods for ROBINS-I risk-of-bias assessment. Two reviewers independently screened, extracted, and cross-checked data. False-alarm rate (FAR) was standardised to /h.

**Results:** Deep learning consistently outperformed conventional machine learning. CNNs and hybrid CNN–recurrent architectures reported up to 99.01% accuracy, 99.81% sensitivity, and 97.70% specificity. Validation was predominantly patient-specific; three studies adopted patient-independent schemes, and none reported cross-dataset external validation. Reporting of prediction horizon was limited, and latency/energy metrics were rarely provided.

**Conclusion:** Despite promising accuracy, the lack of real-world validation and the under-reporting of deployment metrics hinder clinical translation. We recommend standardised evaluation (PS/PI/EXT), an end-to-end latency budget (pre-processing, model runtime, I/O, alert handling), and prospective FAR per 24 h, with prospective and cross-dataset validation to enhance reliability and patient outcomes.

**Highlights:**

- We synthesised 23 real-time EEG seizure-prediction studies (2017–2025).
- Deep learning peaks: Acc 99.01%, Sens 99.81%, Spec 97.70% (not one model).
- Validation was patient-specific; no cross-dataset external validation reported.
- Latency and energy metrics were rarely reported; FAR harmonised to /h.
- Report PS/PI/EXT, end-to-end latency budget, and FAR per 24 h.

## 1 Introduction

Epileptic seizures are neurological events caused by abnormal electrical discharges of neurons from the brain. Epilepsy affects approximately 50 million people worldwide. Individuals with epilepsy may unintentionally harm themselves or others during seizure episodes [1].

Beyond clinical symptoms, due to their sudden, unpredictable, and frequent characteristics, individuals with epilepsy often face long-standing psychosocial barriers, including entrenched stigma and systemic discrimination in cultural and healthcare settings [2, 3].

The clinical potential of real-time seizure prediction is based on its ability to improve patient safety and quality of life. To situate current approaches, we outline three phases—Pre-Digital, Digital, and AI-driven. Each phase provides important contributions, while also exposing its crucial limitations. Table 1 shows details of milestones, the technological features introduced, and how each of them helped inform current approaches to prediction. Performing a timely prediction of precursor seizures can help patients avoid potentially unsafe situations and create a valuable window for clinical intervention.

**Table 1:** Evolution of Epilepsy Management Technologies. Key milestones from pharmacological, surgical, and AI-based systems are presented chronologically to illustrate how each era contributed to real-time seizure prediction readiness.

| Era | Year | Technology Type | Representative Event | Core Limitations | Predictive Insight |
| --- | --- | --- | --- | --- | --- |
| Pre-Digital | 1857–1890s | Pharmacotherapy | Locock’s 1857 report on potassium bromide in 14–15 cases of “hysterical” epilepsy in young women [4] | Non-personalised pharmacodynamics | Highlighted the need for individualised treatment approaches beyond generalised pharmacology |
|  | Late 19th–mid 20th century | Surgical Intervention | Early cortical resections by Horsley and later Penfield [5] | Imprecise localisation; procedural risk | Stimulated the development of EEG-guided localisation methods for accurate seizure focus identification |
| Digital | 2013 | Closed-Loop Neurostimulation | FDA approval of the Responsive Neurostimulation (RNS) system [6] | Bulky hardware; limited power | Demonstrated the feasibility of feedback-driven intervention and emphasised the need for efficient embedded systems. |
|  | 2010s | Multimodal Sensing Integration | Wearable sensors and systems [7, 8] | Synchronisation and heterogeneity issues | Enabled diverse signal acquisition and prompted trade-off analysis between energy efficiency and data fusion quality. |
| AI-Driven | Around 2020s | Multimodal Signal Acquisition | Embedded/edge deployment of multimodal EEG systems (e.g., EEG + ECG fusion with low-power CNNs) [9] | Limited generalisation across subjects | Advanced adaptive modelling and edge computing to support real-time, personalised seizure forecasting. |
|  | 2023–Present | Multimodal Neurocritical Care Data for AI Readiness | Harmonisation initiatives for multimodal neurocritical care data [10] | Regulatory complexity; data protection concerns | Facilitated the convergence of multimodal clinical signals and AI to enable continuous seizure prediction. |

Unlike prior reviews that address a wide range of objectives, the present study focuses specifically on identifying which machine learning (ML) and deep learning (DL) architectures yield the most reliable predictive performance in real-time seizure forecasting. Rather than treating performance metrics in isolation, we assess these models using a layered framework that also considers computational efficiency, energy demands, interpretability, and validation status.

Beyond conventional metrics, deployment-oriented factors (latency, energy cost, interpretability, and validation status) are considered as decision criteria. In this systematic review, we offer an additional assessment framework (Table 2) in conjunction with traditional metrics, including accuracy, sensitivity, specificity, latency, and the practical utility of prediction systems in clinical or wearable settings.

**Table 2:** Evaluation Dimensions for Real-Time Seizure Prediction Models. Supplementary criteria are proposed to assess model viability for deployment in clinical or wearable systems beyond core performance metrics.

| Evaluation Dimension | Scope | Relevance |
| --- | --- | --- |
| Real-time responsiveness | Latency ($\leq 2$ s), runtime, pre-processing time | Determines feasibility for embedded or wearable platforms requiring low-latency computation |
| Interpretability | Attention mechanisms, saliency maps, feature attribution tools | Promotes clinician trust and regulatory transparency for high-stakes clinical applications |
| Multimodal data capacity | EEG-only vs. EEG combined with ECG or motion signals | Improves robustness in variable or noisy recording conditions |

## 2 Methods

### 2.1 Protocol and Registration

The study adhered to the PRISMA guidelines, and the final search was completed on 31 May 2025. The protocol was then registered retrospectively in PROSPERO (CRD420251062076); eligibility criteria, screening procedures, and extraction templates were pre-specified and were not altered after registration. Registration was retrospective because the project began as a scoping review before expanding into a full systematic review; no changes were made to eligibility criteria or analysis plans after registration.

#### 2.2 Sources of Information and Search Date

To ensure thorough retrieval, we systematically searched four major indexing databases: PubMed, IEEE Xplore, Scopus, and ScienceDirect. The search covered publications from 1 January 2017 to 31 May 2025, focusing on studies involving biomedical signal processing and AI-based seizure prediction.

### 2.3 Search Strategy

Eligible records were restricted to peer-reviewed original articles or full-length conference papers. We excluded reviews, non-English reports, retracted items, and preprints from the quantitative synthesis. Search queries were constructed using Boolean logic with the following key terms: “epileptic seizure prediction” OR “seizure forecasting”, AND “machine learning” OR “deep learning” OR “real-time system”, AND (“EEG”[Title/Abstract] OR “ECoG”[Title/Abstract] OR “wearable sensors”[Title/Abstract]). Syntax was tailored to each platform, with IEEE Xplore and ScienceDirect searches restricted to journal articles and full-length conference papers. Table 3 summarises the final queries. Full query strings for each database are provided in Supplementary A.

**Table 3:**
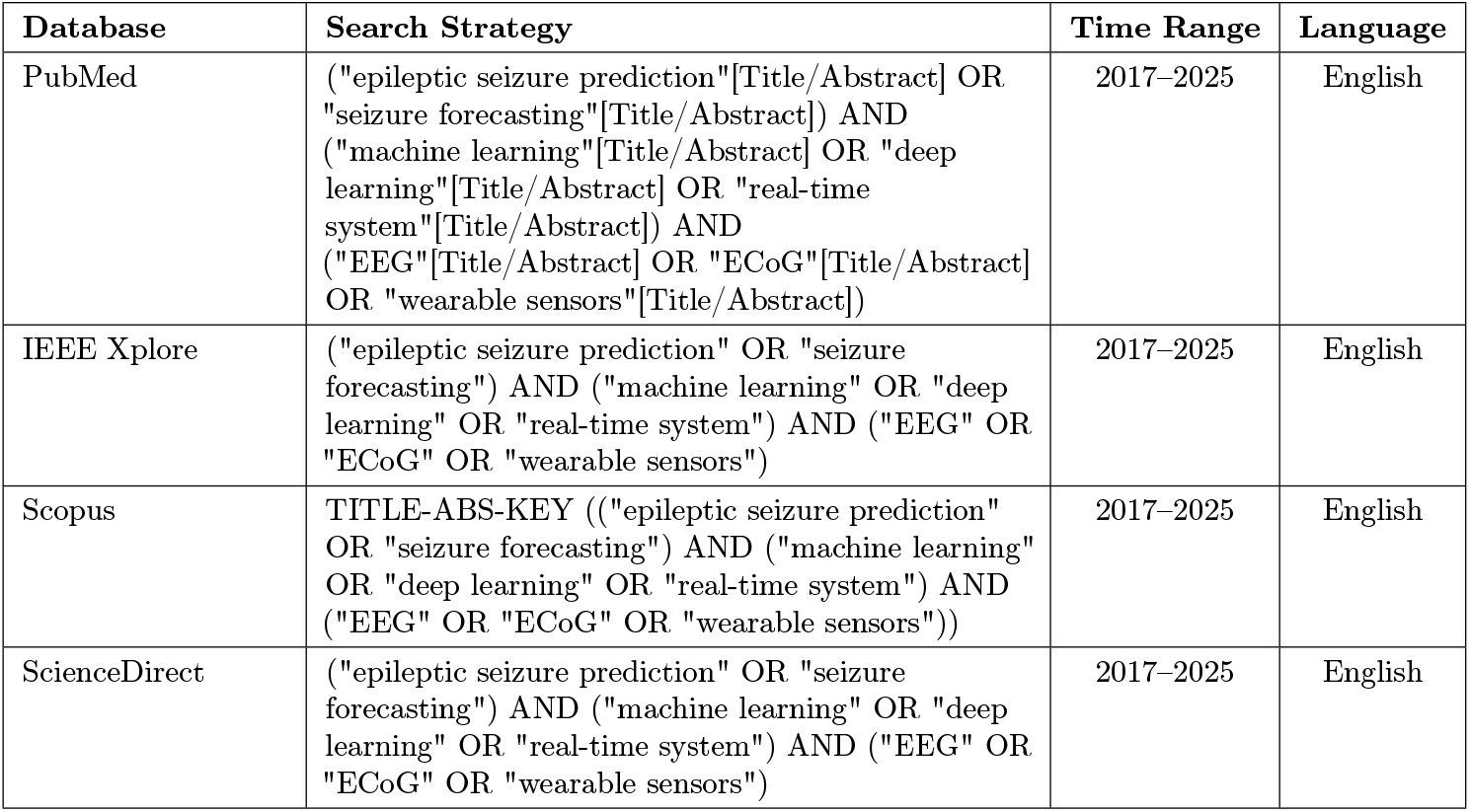
Database-specific Search Strategy for Literature Retrieval.

| Database | Search Strategy | Time Range | Language |
| --- | --- | --- | --- |
| PubMed | ("epileptic seizure prediction"[Title/Abstract] OR "seizure forecasting"[Title/Abstract]) AND ("machine learning"[Title/Abstract] OR "deep learning"[Title/Abstract] OR "real-time system"[Title/Abstract]) AND ("EEG"[Title/Abstract] OR "ECoG"[Title/Abstract] OR "wearable sensors"[Title/Abstract]) | 2017–2025 | English |
| IEEE Xplore | ("epileptic seizure prediction" OR "seizure forecasting") AND ("machine learning" OR "deep learning" OR "real-time system") AND ("EEG" OR "ECoG" OR "wearable sensors") | 2017–2025 | English |
| Scopus | TITLE-ABS-KEY (("epileptic seizure prediction" OR "seizure forecasting") AND ("machine learning" OR "deep learning" OR "real-time system") AND ("EEG" OR "ECoG" OR "wearable sensors")) | 2017–2025 | English |
| ScienceDirect | ("epileptic seizure prediction" OR "seizure forecasting") AND ("machine learning" OR "deep learning" OR "real-time system") AND ("EEG" OR "ECoG" OR "wearable sensors") | 2017–2025 | English |

### 2.4 Study Selection

Two independent reviewers screened titles and abstracts using predefined inclusion and exclusion criteria (Table 4). Records selected by either reviewer were carried forward for full-text evaluation. Screening was performed using the Rayyan platform, and duplicates were removed via Zotero reference manager. Discrepancies were resolved through discussion; unresolved cases were adjudicated by a third reviewer. No machine-learning–based automation was used for study selection; Rayyan was used solely to manage manual screening. When eligibility remained unclear, corresponding authors were contacted for clarification.

**Table 4:** Eligibility Criteria for Inclusion and Exclusion.

|  | Inclusion Criteria | Exclusion Criteria |
| --- | --- | --- |
| **Population / Problem** | Studies involving patients or datasets focused on real-time epileptic seizure prediction | Studies focused solely on diagnosis; studies of non-epileptic conditions |
| **Intervention** | ML/DL models with defined prediction horizon and latency | Offline methods or studies focused only on feature extraction |
| **Comparison** | Benchmarking against other models or baseline methods | No comparative evaluation or single-model reports |
| **Outcomes** | Performance metrics (e.g., accuracy, sensitivity, specificity, F1-score) reported on held-out test sets | No reported performance or absence of validation on independent data |
| **Bias / Analysis** | Sufficient methodological detail enabling ROBINS-I assessment by reviewers (original studies need not provide a risk-of-bias table) | Insufficient methodological detail to permit a ROBINS-I judgement |
| **Study Type** | Peer-reviewed original articles or full-length conference papers | Preprints, abstracts, or reviews without new data |
| **Language and Availability** | English-language, full-text accessible | Non-English or retracted studies; full-text not retrievable even after at least two unsuccessful contact attempts within one month |

### 2.5 Data Collection Processes and Items

Data extraction was conducted by the lead author and validated by a second reviewer. Discrepancies were discussed and resolved, when applicable. Extracted variables included study objectives, dataset characteristics, model type, algorithm architecture, prediction horizon, performance metrics (e.g., sensitivity, latency, false-alarm rate), and validation strategies (cross-validation, external testing). We define validation types as: patient-specific (PS), patient-independent within-dataset (PI), and external cross-dataset/institutional validation (EXT). All false-alarm rates were harmonised to FAR (/h); studies reporting FPR/h were mapped equivalently. Prediction horizon refers to the lead time from alarm to ictal onset. Only figure-generation scripts were used to compute summary statistics from Table 5 and to render the PRISMA diagram, the ROBINS-I plot, and the timeline heatmap; no model training code was executed in this study.

**Table 5:** Integrated model characteristics and evaluation results across seizure prediction studies, including AUC and prediction horizon.

| Study<br>(Author, Year) | Key Findings | Dataset | Valid.<br>(PS/PI/EXT/NR) | Type | Arch. | Acc. (%) | Sens. (%) | Spec. (%) | FAR (/h) | AUC | Hzn (min) |
| --- | --- | --- | --- | --- | --- | --- | --- | --- | --- | --- | --- |
| Parani (2025)[21] | LLM achieved the highest average sensitivity (79.02%) and accuracy (70.80%) across all 12 PI benchmarks. | MLSPred-Bench (PI benchmarks) | PI | DL | Pre-trained LLM (Longformer); base-lines include ViT | 70.80 | 79.02 | 62.53 | NR | 71.40 | 2 / 5 / 15 / 30 |
| He (2023)[2] | Autoencoder-based model achieved **97.32%** sensitivity and 0.009/h FAR without manual feature engineering; AUC 98.44%. | CHB-MIT | PS | AE-based | RTS-RCVAE + Bi-LSTM | 98.43 | **97.32** | NR | 0.009 | 98.44 | NR |
| Lammie (2021)[13] | Memristive DL system (simulated) on CHB-MIT; mean Sens 77.4%, AUROC 0.85; reports per-fold FPR curves; simulation; not end-to-end latency measured. | CHB-MIT | NR | CNN-based | 2D CNN (memristive simulation) | NR | 77.4 | NR | NR | 0.85 | NR |
| Xu (2024)[22] | TSGCN-MLCL with domain adaptation and GAN-based augmentation; reports gains over prior base-lines on CHB-MIT and Kaggle iEEG. | Kaggle iEEG; CHB-MIT | NR | GAN-based | TSGCN-MLCL | NR | NR | NR | NR | NR | NR |
| Li (2023)[14] | Parallel 1D-CNN; CHB-MIT: Acc 99.01%, Sens 99.24%, Spec 98.68%, FAR 0.47/h; SWEC-ETHZ: Acc 97.54%, Sens 98.22%, Spec 97.02%, FAR 0.99/h. | CHB-MIT; SWEC-ETHZ | NR | CNN-based | Parallel 1D-CNN | 99.01 / 97.54 | 99.24 / 98.22 | 98.68 / 97.02 | 0.47 / 0.99 | NR | NR |
| Li (2022)[15] | Memristive CNN achieves 99.01% accuracy on CHB-MIT and 97.54% on SWEC-ETHZ, with 98.68% specificity; designed for efficient low-power in-memory computing. | CHB-MIT, SWEC-ETHZ | NR | CNN-based | Memristive CNN | 99.01 | 99.24 | 98.68 | 0.47 | NR | NR |
| Sadeghzadeh (2019)[3] | Online pre-ictal feature monitoring; Acc 88.76%, Sens 90.62%, Spec 88.34%, FAR 0.0046/h; mean prediction time 23.3 min; single-channel thresholding. | CHB-MIT (19 pts) | PS | Feature-based ML | Line-length + multi-stage thresholds | 88.76 | 90.62 | 88.34 | 0.0046 | NR | 23.3 |
| Varnosfaderani (2024)[26] | Two-layer LSTM using 4 channels; EPILEPSIAE (iEEG European) (26 pts): Acc 78.18%, Sens 85.7%, FAR 0.22/h, AUC 0.885; LOOCV at sample level. | European iEEG | NR | LSTM-based | 2-layer LSTM | 78.18 | 85.7 | NR | 0.22 | 0.885 | 5 |
| Shaik Gadda (2023)[24] | Optimized feature set with XGBoost; leave-one-subject-out evaluation on CHB-MIT. Mean horizon 10.17 min; Acc 83.33%. | CHB-MIT | PI | Feature-based ML | XGBoost (LOSO) | 83.33 | NR | NR | NR | NR | 10.17 |
| Wang and Zhang (2024)[17] | Multiscale convolutional attention network achieved 97.5% sensitivity, FAR 0.30/h, and average warning time 26.04 min on CHB-MIT. | CHB-MIT | NR | CNN-based | MS-CANet | NR | 97.5 | NR | 0.30 | NR | 26.04 |
| Wang (2024)[18] | Multiscale convolutional attention network (MSCAN); Sens 97.5%, FAR 0.30/h; mean prediction time 26.04 min. | CHB-MIT | NR | CNN-based | MSCAN (attention + multiscale) | NR | 97.5 | NR | 0.30 | NR | 26.04 |
| Sarvi Zargar (2023)[25] | MobileNet-V2 and FC achieved 98.39% sensitivity and 0.029/h FAR in PI seizure prediction. | European iEEG (EPILEPSIAE) | PI | CNN-based | Transfer-learning / MobileNet-V2 + FC | NR | 98.39 | NR | 0.029 | NR | 40 |
| Wang (2022)[30] | Residual convolutional VAE with Bi-LSTM achieved 95.94% accuracy, 95.24% sensitivity, FAR 0.015/h, and AUC 96.00% on CHB-MIT (10-fold). | CHB-MIT | PS | AE-based | RCVAE + Bi-LSTM | 95.94 | 95.24 | NR | 0.015 | 96.00 | NR |
| Mustaqeem (2023)[16] | CNN and bandpass filtering model achieved 89.5% accuracy, 89.7% sensitivity, and AUC of 89.5%. Generalized non-patient-specific model. | CHB-MIT | NR | CNN-based | Bandpass CNN | 89.5 | 89.7 | NR | NR | 89.5 | NR |
| Yu (2020)[20] | LMD combined with CNN and Bayesian LDA achieved 87.7% sensitivity and 0.25/h FAR using iEEG from 21 patients. | iEEG (public, 21 pts) | NR | CNN-based | LMD with CNN and LDA | NR | 87.7 | NR | 0.25 | NR | NR |
| Duan (2019)[1] | Multi-Timescale CRNN reached 94.8% accuracy, 91.7% sensitivity, and 97.7% specificity on CHB-MIT. | CHB-MIT | NR | CRNN | Multi-Timescale CRNN | 94.8 | 91.7 | 97.7 | NR | NR | NR |
| Nazari (2022)[31] | Few-shot CNN model achieved 98.52% sensitivity and 0.045/h FAR on CHB-MIT with 5-min horizon. | CHB-MIT (3 pts) | PS (few-shot) | Few-shot CNN | CNN using few-shot learning | NR | 98.52 | NR | 0.045 | NR | 5 |

| Study<br>(Author, Year) | Key<br>Findings | Dataset | Valid. | Type | Arch. | Acc. (%) | Sens. (%) | Spec. (%) | FAR (/h) | AUC | Hzn (min) |
| --- | --- | --- | --- | --- | --- | --- | --- | --- | --- | --- | --- |
| Ntahobari (2022)[28] | SHAP optimized Extra Trees classifier significantly reduces training time ($\times 1.83$) while maintaining seizure prediction accuracy. Zero-crossing is best feature. | Monash and Contest (iEEG) | NR | Feature-based ML | SHAP with Extra Trees | NR | NR | NR | NR | NR | NR |
| Zhao (2022)[29] | NAS with pruning produced energy-efficient model ($< 10$ mJ) with 99.81% sensitivity and 0.005/h FAR on CHB-MIT. Compressed for wearable use. | CHB-MIT, iEEG | PS | CNN-based | NAS with pruned DNN | NR | 99.81 | NR | 0.005 | 1.0 | NR |
| Xu (2020)[19] | End-to-end CNN with 1D and 2D kernels achieved 98.8% sensitivity and 0.074/h FAR on CHB-MIT; outperformed SVM and regression models. | CHB-MIT, iEEG (Kaggle) | NR | CNN-based | End-to-end CNN | NR | 98.8 | NR | 0.074 | 0.988 | NR |
| Varnosfaderani (2024b)[27] | Two-layer LSTM on European iEEG dataset with mean post-processing yielded AUC 0.885; seizure type significantly affects prediction quality. | EPILEPSIAE (iEEG European) | NR | LSTM-based | 2-layer LSTM | NR | NR | NR | NR | 0.885 | NR |
| Yang (2021)[32] | Dual self-attention residual network (RDANet) achieved 92.07% accuracy, 89.33% sensitivity, 93.02% specificity, and AUC 91.26% on CHB-MIT. | CHB-MIT | NR | CNN-based | RDANet | 92.07 | 89.33 | 93.02 | NR | 91.26 | NR |
| Maddineni (2022)[23] | Hybrid transformer framework using FFT and deep transformer achieved 95.2% sensitivity and 0.02/h FAR; robust across EEG conditions. | CHB-MIT | NR | Hybrid DL | FFT and Deep Transformer | NR | 95.2 | NR | 0.02 | NR | NR |

**Notes.** PS = patient-specific; PI = patient-independent (same dataset); EXT = external (cross-dataset or cross-institution). All false-alarm metrics are reported as **FAR (/h)**; where a study reported *FPR/h*, we recorded it as FAR (/h) for consistency. NR = not reported. Li (2022) and Li (2023) report different CNN architectures evaluated on CHB-MIT and SWEC-ETHZ; similar numbers reflect independent reports rather than table duplication. Wang and Zhang (2024) and Wang (2024) report closely related multiscale-convolutional attention variants; we treat them as method variants and do not double-count them in narrative statistics.

### 2.6 Risk of Bias Assessment

Risk of bias was assessed independently by two reviewers using the ROBINS-I tool. Seven domains of bias were considered, each categorised as low, moderate, serious, or critical risk. Overall risk ratings were derived accordingly. Detailed search queries for each database are provided in Supplementary A. Disagreements were resolved by a third reviewer. For prediction modelling, we additionally considered applicability concerns using PROBAST and aligned reporting with TRIPOD where applicable. In addition, PROBAST screening indicated high applicability concerns in the ‘participants’ and ‘analysis’ domains in most studies, consistent with the limited use of PI evaluation and the absence of cross-dataset external validation. We provide domain-level ROBINS-I judgements and brief justifications per study in Supplementary A. Inter-rater reliability for the ROBINS-I assessments across all included studies yielded a mean Cohen’s kappa of 0.56 (range: 0.28–1.00), indicating moderate agreement between the two reviewers according to the Landis and Koch criteria [11]. Discrepancies were resolved through discussion until consensus was reached.

### 2.7 Synthesis Methods

Data synthesis was descriptive. Quantitative metrics were summarised using means or inter-quartile ranges. Meta-analysis was not performed due to high heterogeneity in study designs, model configurations, and validation approaches.

## 3 Results

This section outlines the study selection process, presents key quantitative results, examines heterogeneity by device type and signal modality, and provides the risk-of-bias assessment.

### 3.1 Study Selection

The study selection process is illustrated in Figure 1. The figure shows a Preferred Reporting Items for Systematic Reviews (PRISMA) flow chart, adapted from the PRISMA 2020 guidelines [12]. The initial database search retrieved a total of 586 records. After removing 162 duplicates and ineligible items using Zotero-based de-duplication (see Supplementary B for log), 424 records remained for title and abstract screening. Following this step, 316 articles were excluded for not meeting the relevance criteria, and 108 full-text articles were evaluated for eligibility. Of these, 85 were excluded due to lack of machine learning methodology (n = 51), review/commentary nature (n = 19), or insufficient technical details (n = 15). A total of 23 studies were finally included in the review. PRISMA 2020 flow diagram source and counts per box are provided as editable files.

**Figure 1.**
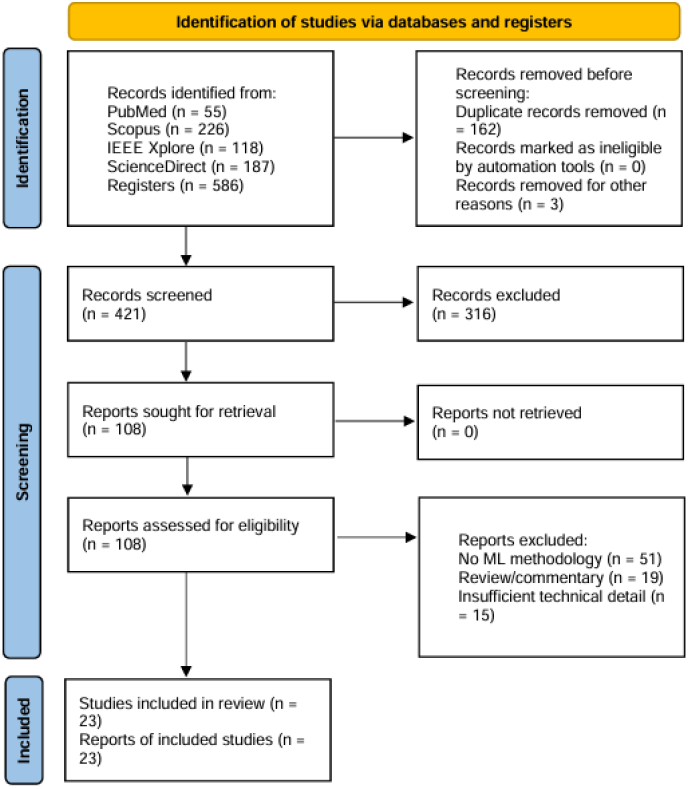
Preferred Reporting Items for Systematic Reviews (PRISMA) flow chart. Duplicates were removed using Zotero; title/abstract screening was conducted in Rayyan.

### 3.2 Study Characteristics

Figure 2 shows the time distribution of the 23 included studies from 2019 to 2025, divided into seven model categories: CNN-based (8 studies), Hybrid DL (6, including LSTM/CRNN hybrids), Transformer-based (3), Feature-based ML (3), GAN-based (1), Few-shot CNN (1), and AE-based (1). In Table 5, validation types are coded as PS (patient-specific), PI (patient-independent, same dataset), and EXT (external, different dataset/institution); false-alarm rate (FAR) is reported per hour (/h); when studies report FPR/h, we treat it equivalently to FAR and harmonise units accordingly.

**Figure 2.**
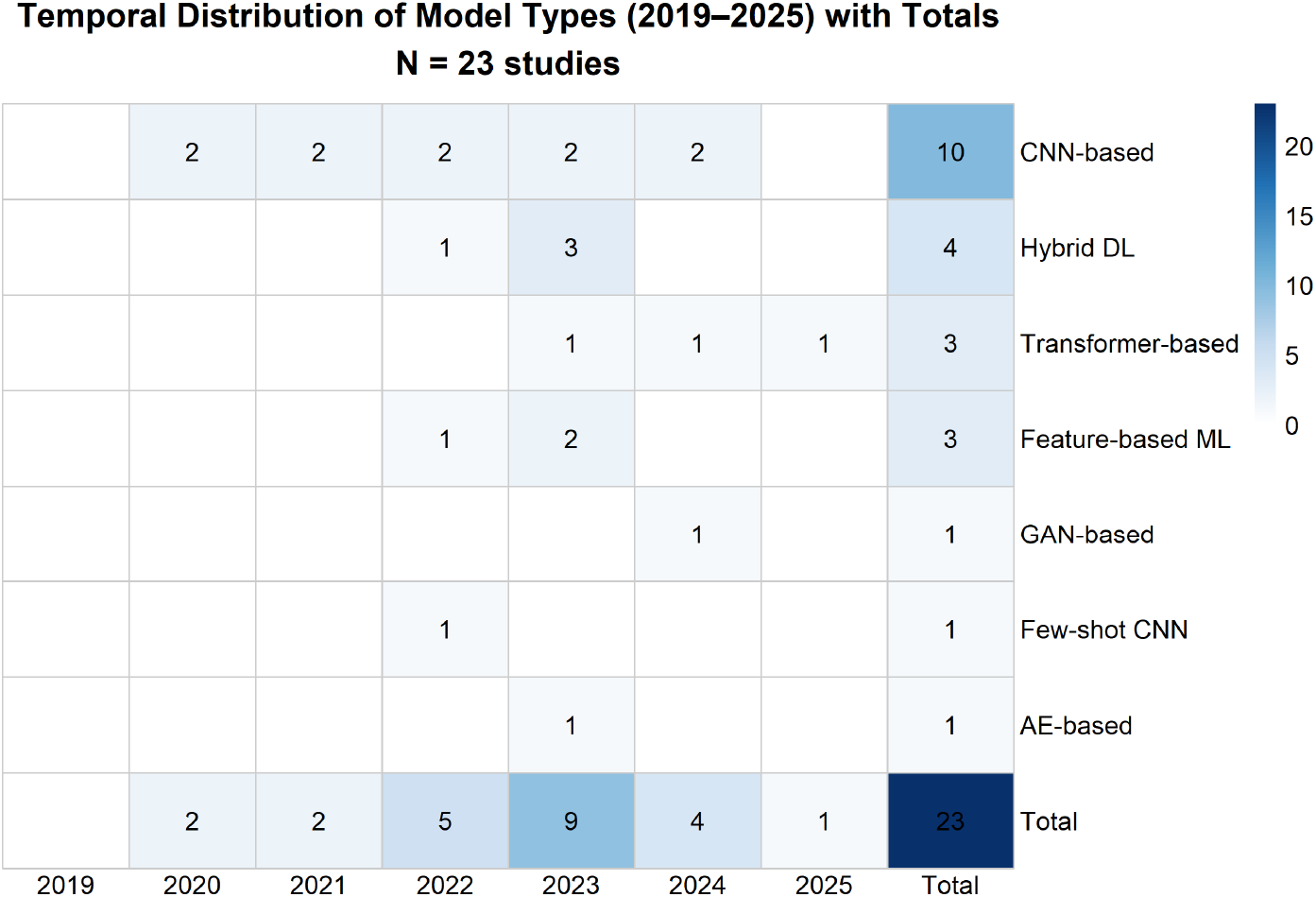
Distribution of included studies by year and model type.

Overall, deep learning approaches appeared to be the most widely used among the 23 studies. Eight studies utilised CNN-based models [13, 14, 15, 16, 17, 18, 19, 20], while others investigated transformer-based [21], GAN-based [22], hybrid transformer frameworks [23], autoencoders [2].

Validation methods varied considerably. The most commonly used internal schemes were 5-fold [14, 15] and 10-fold [1]. A PI, within-dataset strategy using leave-one-subject-out (LOSO) was adopted by [24]. A total of seven studies did not clearly specify their validation scheme. Where unclear, we coded validation as NR and contacted authors when feasible. PI validation was used in three studies [21, 25, 24], and no study reported explicit cross-dataset external validation (see Table 5).

Regarding datasets, CHB-MIT was the most frequently used (17 studies, approximately 74% of the included works), followed by iEEG-derived datasets such as the EPILEPSIAE (European iEEG) [26, 27], Monash iEEG [28], and Kaggle’s AES competition dataset (see Table 5) [19]. Analyses used publicly available or licensed EEG datasets; no included study explicitly reported the use of an unpublished private dataset. Most studies did not explicitly target a specific age group, although the use of CHB-MIT (a paediatric dataset) suggests a focus on younger populations in roughly three-quarters of the works. Only three studies [25, 21, 24] adopted PI evaluation on public datasets; explicit cross-dataset external validation was not reported.

Performance results were reported in a variety of ways. For studies using deep learning (DL) frameworks, sensitivity values reached as high as 99.81% [29]. Interestingly, only eight studies reported the prediction horizon (defined as the lead time between a model’s prediction and the actual onset of a seizure, which is a key measure for enabling proactive clinical intervention).

In terms of study design, 18 of the 23 studies reported using publicly available datasets with ethical approval statements, and all evaluated performance on patient-level EEG segments (segments refer to temporally contiguous portions of EEG recordings, often a few seconds to minutes in length, used as the basic unit for model training and testing). Several studies compared multiple models under identical conditions to allow for fair benchmarking [22, 23]. Across the 23 studies, prediction horizon was explicitly reported in 8 studies; latency was reported in ≤ 2 studies and energy/power in ≤2 studies (where available, metrics were heterogeneous and not end-to-end). No study provided a complete latency budget covering pre-processing, inference, data transfer, and alert handling. We treat closely related multiscale-convolutional attention variants as method variants and do not double-count them in narrative statistics.

Table 5 summarises the model types, datasets, validation approaches, performance metrics, and prediction horizons of all included studies.

### 3.3 Risk of Bias in Studies

As illustrated in Figure 3, D1 (bias due to confounding) had the highest proportion of “serious” risk, with nearly half of the studies judged as serious; the remainder were mostly “moderate,” and none were “low risk.” For D2 (selection of participants), D3 (classification of interventions), D4 (deviations from intended interventions), and D6 (measurement of outcomes), most studies were rated “low risk,” with a small number “moderate” and none “serious.” In D5 (missing data), a substantial proportion were rated “no information,” with the rest “low risk.” In D7 (selection of the reported result), most studies were “moderate,” with the remainder “low risk.” Overall, most studies were judged to have a “moderate” risk of bias, several were “serious,” and no study achieved a “low risk” rating across all domains. Domain-level ROBINS-I judgements and brief justifications are provided in Supplementary Materials, with Figure 3 offering a visual summary.

**Figure 3.**
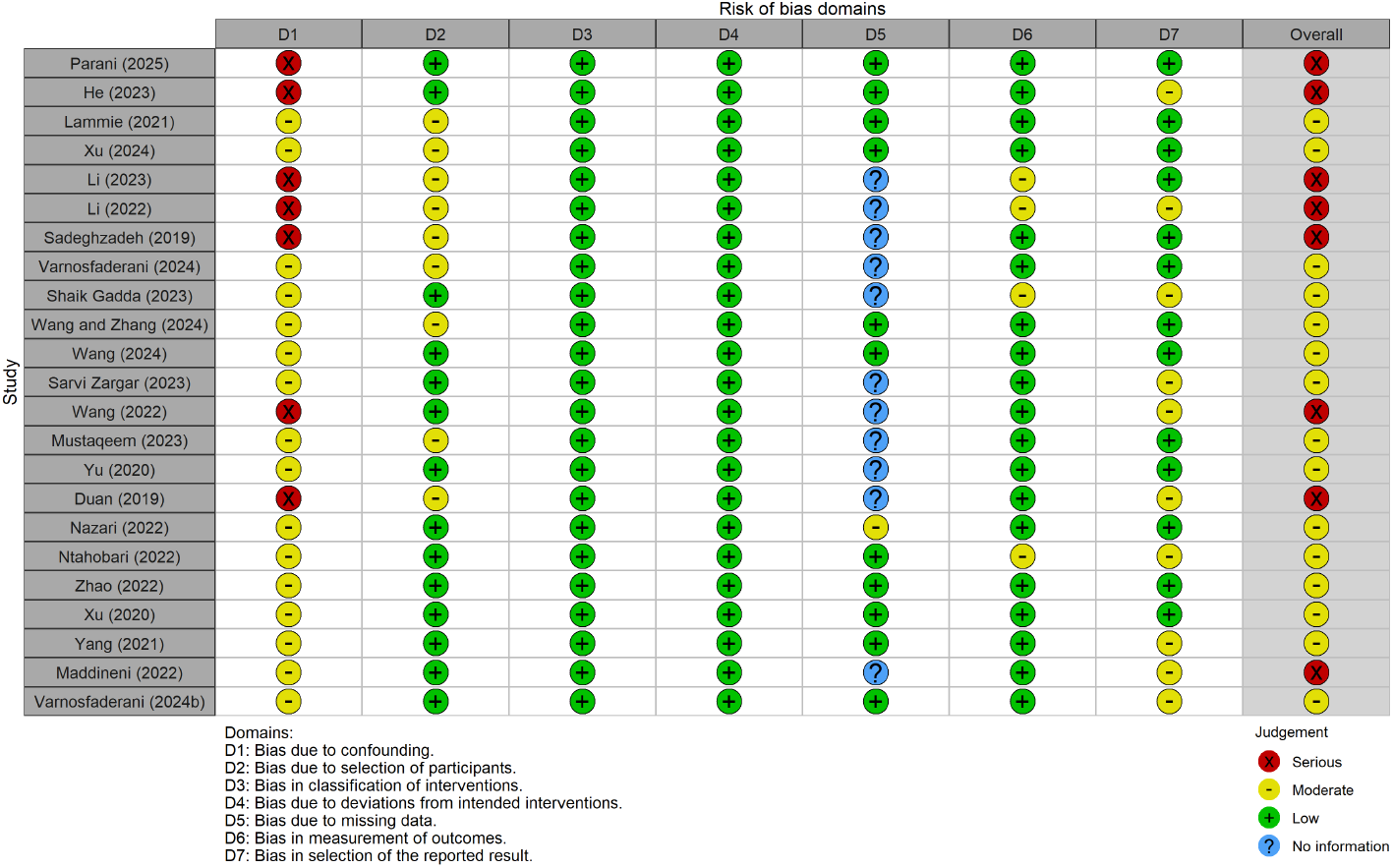
Risk of bias was evaluated across the included non-randomised studies using the Risk Of Bias In Non-randomized Studies of Interventions (ROBINS-I) tool, and the results are summarised in Supplementary Materials.

## 4 Discussion

Electroencephalography (EEG) data has been widely used in seizure prediction and remains the predominant input modality to date. All studies included in this review have used EEG datasets, and the methods employed cover paradigms ranging from traditional machine learning to deep learning. From a usability perspective, EEG spans fixed clinical systems and newer wearable form factors; wearable devices can simplify continuous monitoring and improve adherence, but reduced channel coverage, higher susceptibility to motion and electrode–skin impedance drift can limit reliability in practice. To address the issues revealed by the datasets, the next sections focus on the main challenges and the frameworks currently employed to mitigate them.

### 4.1 Limitations of Dataset Design and Simulation Validity

In recent years, researchers have increasingly emphasized the significant disconnect between standard EEG data collection protocols and actual clinical conditions [33, 34, 35]. The practical applicability of these results remains uncertain outside controlled settings.

For the most widely used CHB-MIT dataset, its acquisition protocol is highly standardised and limited to hospitalized pediatric patients. Furthermore, as shown in Table 6, most of the studies were validated only on CHB-MIT or iEEG datasets, with few conducting cross-dataset evaluations [21, 27]. For instance, Yang et al. [32] reported that a CNN-based model trained on the CHB-MIT dataset exhibited a notable drop in performance when applied to the Monash iEEG corpus, with sensitivity falling from 92.4% to 67.1%. Some studies explored GAN-based approaches, but these were customised to specific datasets and showed limited adaptability [36, 22]. While a few augmentation transformations for EEG data have been proposed in the literature, their positive impact on performance is often evaluated on a single dataset and compared to one or two competing augmentation methods [37]. This highlights the severity of train-test distribution mismatch and its impact on model reliability in real-world deployment. In addition, sample distribution within datasets is often highly imbalanced, with certain patients contributing disproportionately more seizure episodes than others. While some do, the majority of the EEG based machine learning studies do not attempt to assess performance of algorithms across recording sessions or across subjects, instead studies use the whole dataset available for training and testing, using an established k-fold cross validation technique and thus missing performance in a real-life setting on an unseen subject [38]; in particular, leave-one-subject-out (LOSO) strategies are underused.

**Table 6:** Summary of Dataset Characteristics Across the Included Studies.

| Dataset | Seizure Annotation | Medication Record | Recording Duration | Demographic Coverage | Annotation Format | Artifact Annotation | Public Availability |
| --- | --- | --- | --- | --- | --- | --- | --- |
| CHB-MIT | No | No | Long | Children | Standardised | Limited | Yes |
| EPILEPSIAE<br>(iEEG European),<br>Monash, Kaggle | Yes | Rarely | Medium to Long | Mixed | Partial | Present | Partially |
| SWEC-ETHZ | Unknown | No | Short | Unknown | Sparse | None | Yes |
| Bonn | No | No | Very Short | Adult | Structured | Minimal | Yes |
| Private (Clinical) | Yes | Unknown | Long | Varied | Not Disclosed | Uncertain | No |

This idealised dataset usually lacks key clinical metadata, including intervention measures and the timing of drug administration. Furthermore, artifacts caused by motion and electrophysiological interferences are usually absent or underrepresented, further distancing model training from the constraints of the real world. Therefore, the models developed under such conditions are difficult to be extended to the demand for real-time and context-aware epilepsy prediction.

While many studies report high classification accuracy, the clinical efficacy and real-world reliability of these models remain in doubt [39, 40]. The core issue is that an epileptic seizure is not an isolated event. On the contrary, it is dynamically influenced by the individual’s physiological state, the external environment and the ongoing clinical background. Therefore, even if the models perform very well on idealized datasets, they are prone to substantial performance degradation in clinical deployment.

The annotation methods of epileptic seizure types are obviously inconsistent across different datasets. Some datasets used expert manual labeling of seizure onset and offset, and some datasets used algorithmic identification[41]. At the same time, outcome labeling results were not uniform, with differences between multicategorical and dichotomous labels. In this systematic review, a total of six EEG datasets were involved, among which the most commonly used were CHB-MIT (17 studies), iEEG-derived datasets including European Epilepsia, Monash, and Kaggle cohorts (6 studies), SWEC-ETHZ(1 study), and Bonn (1 study). Furthermore, A few studies additionally referenced local clinical cohorts; however, primary analyses were conducted on public datasets. Due to the strong openness of CHB-MIT, along with its high signal quality and standardised format, it has become the first choice for most researchers. However, all the signals in this dataset originate from children and lack key information such as seizure types and medication records, raising concerns about its generalisability [33]. The Bonn dataset provides clean data and strong label consistency. Compared with CHB-MIT, the Bonn dataset presents a clearer structure and more consistent annotations, although it has its own limitations. However, the data fragments are very short and are usually used for static classification tasks rather than real-time prediction. Therefore, models trained on this dataset often report very good results, but such performance may be seriously overestimated [42]. Some non-public institutional cohorts described in the literature may reflect more realistic clinical conditions; however, because these data are not publicly accessible, reuse and cross-study comparison are limited. In our included set, primary analyses were conducted on public datasets. Beyond annotation protocols, terminological inconsistencies also complicate evaluation and interpretation. In this review, we adopt the following working definitions for consistency: a seizure segment refers to a short EEG excerpt centered on the ictal peak, often lasting less than 30 seconds and used primarily for classification tasks; a seizure event represents the entire process of an epileptic seizure, from before the seizure to recovery after it. The inconsistent use of them may lead to differences in performance indicators such as delay, sensitivity or false alarm rate.

Currently, the publicly available epilepsy electroencephalogram (EEG) data sets generally have biases in sampling strategies. On the one hand, many studies tend to select datasets with a higher citation volume for modelling and evaluation, especially CHB-MIT. This research orientation can lead to selective bias, especially in these datasets where the test results themselves tend to be overly optimistic and deviate from reality [39]. For example, some highly cited papers have adopted idealized samples with simple structure, less interference and high degree of standardisation in the annotations [39]. Their verification scenarios are far from the complexity of real clinical practice. These results lack reference value to a certain extent. On the other hand, when researchers use datasets with small sample sizes or relatively simple sampling designs, the model performance metrics often show significant fluctuations. These limitations pose significant challenges to the generalisability of EEG-based prediction models and call for careful dataset design and standardised labelling protocols in future work. Without addressing these fundamental issues, the reliability and clinical applicability of seizure prediction models will remain severely constrained.

### 4.2 Real-Time Implementation Challenges: Computational Latency, Hardware Limitations, and Signal Integrity Issues

Real-time seizure prediction faces challenges arising from human factors and practical constraints, including system latency, limited hardware performance, and signal noise. These factors affect the development and application of real-time systems.

Scalp EEG has a low signal-to-noise ratio (SNR). Accurate prediction requires both temporal and spatial information, and reduced SNR can constrain performance, especially outside controlled settings. Recently, several studies have attempted to solve this problem from different aspects, but the current solutions have not reached a consensus. Li et al. [15] proposed a novel low-latency parallel CNN architecture and conducted 5-fold cross-validation on the Bonn, CHB-MIT, and SWEC-ETHZ datasets. The architectures achieved 99.01 % and 97.54 % in seizure prediction, respectively. In simulations, Lammie et al. [13] evaluated the feasibility of using the Memristor Deep Learning System (MDLS) for real-time seizure prediction on the edge side. Under the MemTorch simulation framework and in combination with the Boston Children’s Hospital–MIT (CHB-MIT) dataset, various MDLS configurations were tested. The results show that under the 65 nm CMOS process, the optimal configuration for processing the EEG spectrogram containing 7,680 samples has a delay of 1.408 ms, a power consumption of 0.0133 W, and a footprint of only 0.1269 mm^2^. The system achieved an average sensitivity of 77.4 % and an area under the ROC curve (AUROC) of 0.85. The MDLS figures (1.408 ms; 0.0133 W; 65 nm CMOS) are simulation-level estimates, not measurements from a fabricated chip.

Existing research has proved that low latency can be achieved through lightweight image models or memristor parallel structures. In edge computing platforms or embedded environments, low latency and power consumption can be achieved, but often at the expense of sensitivity. Therefore, the current limitation is that no recognised solution exists to solve this problem. This indicates that the latency issue is not merely an engineering challenge, but stems from the inherent structure of most DL models that rely on full-window inputs. Moreover, while summarising the results, it was found that most studies paid more attention to results such as sensitivity and accuracy, and real-time performance metrics were generally under-reported. Furthermore, the absence of standardised latency-evaluation protocols likely contributes to under-reporting.

As mentioned earlier, the evaluation criteria for latency are often restricted by hardware development. Not only in this aspect, but with hardware advancements, many limiting problems could also be effectively addressed. Most deep learning models studied need to be deployed on high-power-consuming GPUs due to excessive computational demands of these models. Therefore, many studies have attempted to solve this problem. Li et al. [15] proposed a “stuck-weight offset” method to alleviate performance degradation caused by the stuck R_ON_/R_OFF_ weights of the memristor, which can restore accuracy by up to 32% without retraining. The CNN module of their platform consumes approximately 2.791 W and occupies an area of 31.255 mm^2^ under the 22 nm FDSOI CMOS process. Werner et al. [43] proposed a highly energy-efficient seizure detection method involving TC-ResNet and time-series analysis. The average power consumption measured using the low-power AI accelerator UltraTrail is only 495 nW. However, energy conservation often comes at the cost of sensitivity and real-time performance. Although most studies have reported excellent results, the lack of verification on mainstream wearable platforms may limit system-level integration. Meanwhile, the absence of standardised evaluation protocols also constrains the balance between performance and practical deployment.

EEG signals convey important information about brain activity in healthy and pathological conditions. However, they are essentially noisy, which poses significant challenges for accurate analysis and interpretation. Kilicarslan et al. proposed a robust adaptive denoising framework, which characterizes and processes motion artifact contamination in EEG measurements using nonlinear mapping based on Volterra [44]. Aquilue-Llorens & Soria-Frisch proposed LSTEEG, a novel LSTM-based autoencoder specifically for artifact detection and correction in EEG signals [45]. From traditional filtering methods to advanced LSTM results, the performance effect has gradually improved, and the methods are gradually advancing. However, most of the methods show that the results are valid, but they have not yet been applied in real-time systems, so the effect is questionable. For wearable-targeted deployments, latency budgets must account for on-sensor pre-processing, data transmission, and edge inference in addition to the model’s computation time. These components are rarely reported as an end-to-end figure, which complicates comparisons across studies and obscures practical readiness.

### Barriers to Clinical Integration

Machine learning algorithms for seizure prediction have shown considerable diagnostic potential, with reported accuracies reaching up to 100%. However, only a few published algorithms have fully addressed the requirements necessary for successful clinical translation. This is largely due to the fact that the properties of training data may limit the generalisability of algorithms; for instance, algorithm performance may vary significantly depending on which EEG acquisition hardware is used, or on differences in electrode placement and patient demographics. Furthermore, the runtime processing cost can be prohibitive in real-time clinical applications, especially in embedded or wearable environments with limited latency and computing resources. A critical assessment of the potential practical effectiveness of machine learning algorithms can help accelerate clinical translation and identify gaps in current epilepsy detection literature. Without a strict and context-aware verification framework, the high-tech performance under simulation conditions will still be insufficient for safe and effective clinical deployment. In practice, integration requires end-to-end workflows (latency budget, alarm policies, and human-in-the-loop review) rather than model-only metrics.

A practical consideration is the growing use of wearable EEG for ambulatory monitoring. These devices can lower setup burden and support long-term use outside clinics, which directly improves accessibility and user adherence. However, current wearable configurations typically operate with fewer electrodes, less stable contact impedance (especially with dry or semi-dry sensors), and higher exposure to motion and environmental artifacts. These factors reduce spatial resolution and degrade signal fidelity, which in turn increases uncertainty in event prediction and can inflate false alarm rates. Moreover, end-to-end system constraints—battery capacity, on-device inference budgets, wireless transmission overhead, and the need for periodic recalibration—can interact with algorithm design choices. Consequently, while wearables may make seizure prediction easier to use, they do not automatically make it more reliable without advances in artifact suppression, adaptive calibration, and validation on data acquired with wearable hardware.

### Lack of External Validation

The lack of rigorous external validation significantly hinders the adoption of seizure prediction systems in clinical settings. Many models demonstrate high accuracy on internal datasets but fail to maintain performance when applied to external, heterogeneous data sources. While most methods report high accuracies on benchmark datasets, they are rarely evaluated on external cohorts, which limits generalisability.

Furthermore, most validation studies were conducted on pre-segmented or retrospectively labelled electroencephalogram (EEG) data, which does not reflect the challenges of continuous and prospective data streams in clinical practice. Even models that perform well on retrospective data may have difficulty handling noisy, artificial or unbalanced real-time electroencephalogram (EEG) signals. This difference has raised concerns about the operational reliability of many proposed models outside laboratory conditions. As shown in Table 5, among the 23 studies included in this review, [21, 25] applied PI validation (PI, across subjects within the same dataset). Clear cross-dataset external validation (EXT) was not explicitly documented. Most relied on internal validation strategies, such as k-fold cross-validation or keeping one subject. Seven studies did not explicitly report their validation methods, reflecting the persistent problem of methodological transparency.

Efforts to solve this problem include formulating an evaluation framework. Dan et al. proposed the SzCORE (Epilepsy Predictive Clinical Outcomes and Real-world Assessment) framework to standardise benchmark protocols and assess clinical readiness. This framework emphasizes the use of consistent assessment methods, clinically relevant performance indicators, and independent patient cohorts [46].

Furthermore, in specific subpopulations, universal problems have been proved. Tapani et al. evaluated the support vector machine-based detector using an external dataset of 28 newborns and demonstrated that “diverse training data is crucial for ensuring performance comparable to that of human experts” [41]. Similarly, Yang et al. reported that “artificial intelligence systems trained on data from a single continent showed significant performance degradation when testing electroencephalograms from different geographical regions”. The importance of training in populationally and geographically different cohorts [32] was emphasized.

Overall, these findings emphasize the need for external validation on various real EEG datasets. Without such verification, the performance of the reported epilepsy detection algorithm may not be reliably translated into clinical practice. These patterns are consistent with the ROBINS-I findings, where confounding (D1) and selection of the reported result (D7) contributed to non-low overall risk, underscoring the need for prospective PI/EXT evaluation.

### Discussion of the Solutions and Methods

The prediction of epileptic seizures using EEG data involves a pipeline that includes data acquisition, pre-processing, and classification, as shown in Figure 4. Each stage introduces specific technical approaches and challenges.

**Figure 4.**
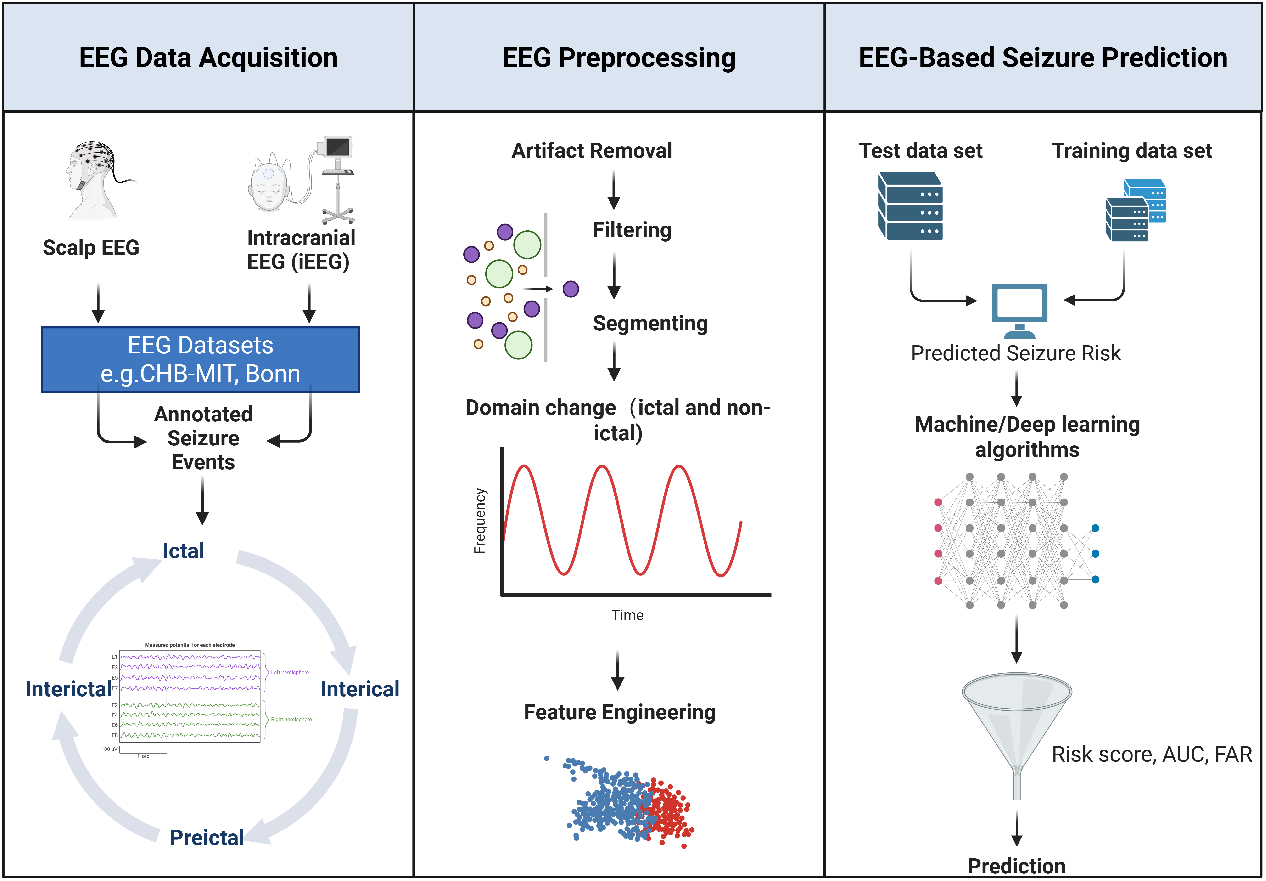
Overview of the EEG-based seizure prediction pipeline, including data acquisition, pre-processing, and model-based prediction.

The first step is to collect data on brain activity. Commonly used techniques include scalp EEG and intracranial EEG (iEEG). The electrode is fixed to the scalp with adhesive and placed according to the international 10–20 system. These electrodes are connected to an electroencephalogram (EEG) device, which records changes in potential as well as spatial and temporal information. The recorded signals were reviewed by experts and classified into epileptic and non-epileptic states. Public datasets such as CHB-MIT and Bonn provide annotated EEG samples widely used for model training and evaluation.

After the EEG data is collected, pre-processing must be carried out. This stage includes removing artifacts, filtering and segmentation. Signals are usually transformed from the time domain to the frequency or time-frequency domain. This step enhances the model’s ability to capture spectral patterns related to epileptic seizures. Then, feature engineering converts the cleaned signals into structured inputs suitable for machine learning. This involves aligning data by class and extracting class-specific attributes.

Different methods are used to extract predictive features. Traditional machine learning methods rely on time-domain features such as entropy and amplitude. These features have computational efficiency and interpretability. However, they may not be able to capture the nonlinear dynamics of epileptic seizure patterns fully. Deep learning models offer another option. Models like CNN and RNN can learn temporal and spectral patterns from raw or transformed electroencephalogram (EEG) signals. Timefrequency representations such as STFT and wavelet transform are usually inputs, enabling CNN to extract hierarchical features. Attention-based hybrid architectures such as CNN-LSTM and Transformers have improved prediction performance. For example, Xu et al. [22] adopted a GAN-based temporal spectrum enhancement scheme, which increased the model sensitivity of the same dataset from 74.2% to 81.6%. However, these improvements cannot be consistently converted between datasets, which indicates limited cross-domain portability. Meanwhile, hybrid models demonstrated relatively balanced results across all metrics.

Despite these advancements, there are still some limitations. Many models are trained and tested on PS data, which limits their universality. The variability of electrode placement, sampling frequency and noise level of the data set reduces reproducibility. The limited availability of predictive data leads to class imbalance, making training more difficult. Although enhancement methods such as GAN and synthetic EEG generation have been explored, their effectiveness depends on the specific datasets and models used.

In terms of deployment, deep learning models, while achieving impressive accuracy, are often computationally intensive, constraining their feasibility for real-time or portable applications. We recommend reporting a latency budget comprising pre-processing, windowing, model inference, data transfer, and alert handling; end-to-end latency should be benchmarked on target hardware. To address this, lightweight architectures such as EEGNet have been proposed. EEGNet, based on depthwise separable convolutions, has demonstrated generalisability across different BCI paradigms, though its robustness in cross-task settings remains uncertain. Quantisation and pruning techniques, such as those used in Q-EEGNet, have also been introduced to reduce computational load, achieving fast inference with minimal energy consumption. However, these methods may compromise performance under noisy or unstructured EEG conditions.

Interpretability also remains a critical concern. Although methods such as saliency maps, SHAP values, and DeepLIFT have been applied to visualise model decisions, their clinical utility is still limited. Deep learning models are often viewed as “black boxes” by clinicians, which undermines trust and hinders practical adoption. We suggest linking explanations to clinically meaningful EEG attributes (e.g., channel-level contributions, frequency bands) and quantifying explanation stability across sessions.

In conclusion, current EEG-based seizure prediction models represent a trade-off between accuracy, computational efficiency, and interpretability. Future work should enhance model generalisation, optimise lightweight architectures for edge deployment, and develop clinically meaningful interpretability tools to facilitate clinical integration.

#### Research gaps and immediate priorities

- **External validation (EXT):** evaluate on an independent dataset/institution; prefer prospective PI/EXT cohorts; report data-shift characteristics.
- **End-to-end latency budget:** provide a hardware-specific budget (pre-processing, windowing, inference, I/O, alert handling) and energy/power; target total latency ≤ 2 s where clinically appropriate.
- **Prospective false-alarm burden:** report FAR per 24 h with refractory policies and seizure-free time; include uncertainty (e.g., 95% CIs).

### 4.3 Conclusion and Outlook

This review evaluated 23 studies on real-time EEG-based epileptic seizure prediction. CNNs and hybrid models integrating recurrent architectures such as LSTM or GRU consistently outperformed classical ML methods regarding sensitivity and specificity. Nevertheless, challenges remain—most notably, dataset heterogeneity, limited real-time validation, and insufficient reporting of system-level performance indicators.

To enable clinical translation, future work should emphasise:

- Standardise dataset curation and evaluation reports (PS/PI/EXT, latency budget, FAR per 24 h).
- Conduct prospective studies and cross-dataset external validation to establish generalisability.
- Integrate alerts into clinical workflows with human-in-the-loop review and refractory policies.
- Advance personalised models while preserving reproducibility and auditability through transparent code and reporting.

Addressing these areas will facilitate the transition from research prototypes to clinically deployable EEG-based seizure prediction systems that can improve patient outcomes.

## Supporting information

Supplementary Materials (ROBINS-I assessment tables and dataset summary)

## ACKNOWLEDGEMENTS

We thank study participants for contributing their time for this study. Along with Rhiannon Walton who coordinated a majority of the study sessions, we thank study team members Samantha LaPorta and Christina Marsicano for their additional assistance with recruitment and data collection. We thank the Medical University of South Carolina’s Alcohol Research Center – and especially Drs. Howard Becker, Patrick Mulholland, Jim Prisciandaro, and William Mellick – for supporting this research.

## Declarations

### Data availability

All primary datasets discussed are public or accessible under license (e.g., CHB-MIT, Bonn, SWEC-ETHZ, European iEEG/EPILEPSIAE, Kaggle AES). Access requirements vary by resource; exact sources, landing pages, and any license notes are listed in Table 5 and the Supplement. No new data were generated.

### Code availability

This systematic review used code solely for figure generation and descriptive statistics; no model training or algorithm development was performed. The scripts reproduce the figures in this article (e.g., PRISMA flow diagram, risk-of-bias plot based on ROBINS-I, and the study timeline heatmap) from the extracted table values. The code will be made publicly available upon publication via Zenodo (DOI to be issued then) and mirrored on GitHub. During peer review, the scripts are available from the corresponding author upon reasonable request.

### Use of generative AI in writing

Language polishing and consistency checks were assisted by a generative AI tool under human oversight; the authors reviewed and are responsible for the content. The tool was used solely for language consistency under human oversight; it was not used for evidence identification, data extraction, or analysis.

### Ethical approval

Not applicable; this study is a systematic review of publicly available reports.

