## Supplementary Materials (ROBINS-I assessment tables and dataset summary) for "Real-Time EEG-Based Epileptic Seizure Prediction Using Artificial Intelligence: A Systematic Review"

### Supplementary A: Risk of Bias Assessment using ROBINS-I

This appendix presents the detailed risk of bias assessment for the 23 included studies, based on the ROBINS-I tool. Each study was independently assessed by two reviewers (Zikang and Kim) across seven bias domains, and an overall judgement was assigned according to the ROBINS-I guidance.

Inter-rater agreement was evaluated using Cohen's Kappa statistic. In cases where discrepancies occurred between the two reviewers, especially when domain ratings differed in severity, consensus was reached through discussion with reference to the ROBINS-I criteria. The final ratings reflect the outcome of this consensus process. Possible ratings include Low risk, Moderate risk, Serious risk, Critical risk, or No Information.

Table 1: *
Risk of Bias Assessment — Parani (2025)[1]

| **Domain** | **Judgement** | **Justification** |
|---|---|---|
| Bias due to confounding | Serious | No confounders described or adjusted for. |
| Bias in selection of participants | Low | Patient-independent MLSPred-Bench used. |
| Bias in classification of interventions | Low | ViT-1, ViT-2, LLM, and ResNet clearly described. |
| Bias due to deviations from intended interventions | Low | No deviations reported; procedures followed. |
| Bias due to missing data | Low | No missing data described or implied. |
| Bias in measurement of outcomes | Low | Accuracy, sensitivity, and specificity all reported. |
| Bias in selection of the reported result | Low | All 12 benchmark results reported for all models. |
| **Overall Risk of Bias** | **Serious** | Cohen's Kappa = 0.46 |

Table 2: *
Risk of Bias Assessment — He (2023)[2]

| **Domain** | **Judgement** | **Justification** |
|---|---|---|
| Bias due to confounding | Serious | No patient-level variables described or adjusted for. |
| Bias in selection of participants | Low | All CHB-MIT subjects used with fixed preictal windows. |
| Bias in classification of interventions | Low | Residual VAE architecture clearly defined. |
| Bias due to deviations from intended interventions | Low | No deviation from described method. |
| Bias due to missing data | Low | No mention of missing or excluded EEG data. |
| Bias in measurement of outcomes | Low | Accuracy, Sensitivity, FPR, AUC reported on test data. |
| Bias in selection of the reported result | Moderate | No registration or reporting protocol described. |
| **Overall Risk of Bias** | **Serious** | Cohen's Kappa = 0.61 |

Table 3: *
Risk of Bias Assessment — Lammie (2021)[3]

| **Domain** | **Judgement** | **Justification** |
|---|---|---|
| Bias due to confounding | Moderate | Patient data in dataset well described however no adjustors described. |
| Bias in selection of participants | Moderate | Only five patients from CHB-MIT dataset used and no reason provided. |
| Bias in classification of interventions | Low | Memristive DL architecture and simulation pipeline fully described. |
| Bias due to deviations from intended interventions | Low | No deviations from stated protocol reported. |
| Bias due to missing data | Low | No missing data described. |
| Bias in measurement of outcomes | Low | Accuracy, sensitivity, AUROC, and the FPR all measured. |
| Bias in selection of the reported result | Low | All reported results cover every tested configuration; no sign of selective omission. |
| **Overall Risk of Bias** | **Moderate** | Cohen's Kappa = 0.30 |

Table 4: *
Risk of Bias Assessment — Xu (2024)[4]

| **Domain** | **Judgement** | **Justification** |
|---|---|---|
| Bias due to confounding | Moderate | No demographic or seizure-type adjustment. |
| Bias in selection of participants | Moderate | 19 / 23 CHB-MIT patients; exclusion unexplained. |
| Bias in classification of interventions | Low | TS-GAN + MLCL architecture fully documented. |
| Bias due to deviations from intended interventions | Low | No protocol deviations reported. |
| Bias due to missing data | Low | Paper reports no missing EEG segments. |
| Bias in measurement of outcomes | Low | Sensitivity, AUC, FPR stated for held-out data. |
| Bias in selection of the reported result | Low | All primary results and comparisons presented. |
| **Overall Risk of Bias** | **Moderate** | Cohen's Kappa = 0.59 |

Table 5: *
Risk of Bias Assessment — Li (2023)[5]

| **Domain** | **Judgement** | **Justification** |
|---|---|---|
| Bias due to confounding | Serious | No patient-level data or confounder adjustment. |
| Bias in selection of participants | Moderate | Public datasets mentioned, cohort coverage unclear. |
| Bias in classification of interventions | Low | Parallel memristive CNN architecture fully described. |
| Bias due to deviations from intended interventions | Low | No reported deviations from planned simulation. |
| Bias due to missing data | No Information | Paper gives no statement on missing EEG segments. |
| Bias in measurement of outcomes | Moderate | Clinical metrics limited; Sens / Spec not comprehensive. |
| Bias in selection of the reported result | Low | All listed metrics reported in Table 2. |
| **Overall Risk of Bias** | **Serious** | Cohen's Kappa = 0.79 |

Table 6: *
Risk of Bias Assessment — Li (2022)[6]

| Domain | Judgement | Justification |
|---|---|---|
| Bias due to confounding | Serious | No patient demographics or confounder adjustment. |
| Bias in selection of participants | Moderate | Three public datasets cited; cohort coverage unclear. |
| Bias in classification of interventions | Low | Parallel memristive CNN pipeline fully documented. |
| Bias due to deviations from intended interventions | Low | No deviations from planned simulation reported. |
| Bias due to missing data | No Information | Handling of missing / corrupted EEG not described. |
| Bias in measurement of outcomes | Moderate | Sens, Spec, Acc, FPR given; clinical breadth limited. |
| Bias in selection of the reported result | Moderate | Results shown for selected configs; no preregistration. |
| **Overall Risk of Bias** | **Serious** | Cohen's Kappa = 1.00 |

Table 7: *
Risk of Bias Assessment — Sadeghzadeh (2019)[7]

| Domain | Judgement | Justification |
|---|---|---|
| Bias due to confounding | Serious | Single-centre CHB-MIT cohort; no demographic or seizure-type adjustment. |
| Bias in selection of participants | Moderate | N = 19/23 CHB-MIT patients; inclusion rationale not detailed. |
| Bias in classification of interventions | Low | Three-level feature-extraction pipeline fully documented. |
| Bias due to deviations from intended interventions | Low | Algorithm executed as stated; no deviations reported. |
| Bias due to missing data | No Information | Paper gives no statement on missing / excluded EEG segments. |
| Bias in measurement of outcomes | Low | Sensitivity, Specificity, Accuracy, FPR all reported on held-out data. |
| Bias in selection of the reported result | Low | All primary metrics presented; no evidence of selective reporting. |
| **Overall Risk of Bias** | **Serious** | Cohen's Kappa = 0.68 |

Table 8: *
Risk of Bias Assessment — Varnosfaderani (2024)[8]

| Domain | Judgement | Justification |
|---|---|---|
| Bias due to confounding | Moderate | Dataset demographics reported, but no covariate adjustment in modelling. |
| Bias in selection of participants | Moderate | 26 / 30 iEEG-Europe patients analysed; exclusion rationale not explained. |
| Bias in classification of interventions | Low | Two-layer LSTM architecture and feature sets described in detail. |
| Bias due to deviations from intended interventions | Low | Pipeline executed as planned; no deviations noted. |
| Bias due to missing data | No Information | Paper gives no statement on missing / excluded segments or artefacts. |
| Bias in measurement of outcomes | Low | Sensitivity, FPR, Accuracy, AUC reported on LOOCV; objective metrics. |
| Bias in selection of the reported result | Low | All primary metrics shown for each patient; no evidence of selective reporting. |
| **Overall Risk of Bias** | **Moderate** | Cohen's Kappa = 0.30 |

Table 9: *
Risk of Bias Assessment — Shaik Gadda (2023)[9]

| Domain | Judgement | Justification |
|---|---|---|
| Bias due to confounding | Moderate | Public dataset; no demographic or seizure-type adjustment. |
| Bias in selection of participants | Low | 22 / 24 CHB-MIT subjects used; two exclusions explained. |
| Bias in classification of interventions | Low | XGBoost and feature-extraction pipeline clearly documented. |
| Bias due to deviations from intended interventions | Low | Model executed as planned; no deviations reported. |
| Bias due to missing data | No Information | Paper does not state how missing / artefact segments were handled. |
| Bias in measurement of outcomes | Moderate | Accuracy, recall score, specificity, and F-measure were measured however minimal detail provided. |
| Bias in selection of the reported result | Moderate | Emphasis on best interval; other diagnostics summarised briefly. |
| **Overall Risk of Bias** | **Moderate** | Cohen's Kappa = 0.28 |

Table 10: *
Risk of Bias Assessment — Wang and Zhang (2024)[10]

| **Domain** | **Judgement** | **Justification** |
|---|---|---|
| Bias due to confounding | Moderate | No demographic/seizure-type adjustment; single benchmark only. |
| Bias in selection of participants | Moderate | First 10 of 24 CHB-MIT subjects analysed; exclusion rationale not provided. |
| Bias in classification of interventions | Low | Multiscale-convolution and attention modules documented in detail. |
| Bias due to deviations from intended interventions | Low | Training/evaluation followed stated protocol; no deviations noted. |
| Bias due to missing data | Low | Authors state no missing EEG segments in selected cohort. |
| Bias in measurement of outcomes | Low | Sens, FPR, PT, Acc reported with clear SPH/SOP definition. |
| Bias in selection of the reported result | Low | Metrics for all subjects and all experiments (BM1-BM12) fully presented. |
| **Overall Risk of Bias** | **Moderate** | Cohen's Kappa = 0.30 |

Table 11: *
Risk of Bias Assessment — Wang (2024)[11]

| **Domain** | **Judgement** | **Justification** |
|---|---|---|
| Bias due to confounding | Moderate | Patients details not described however used cross-validation . |
| Bias in selection of participants | Low | CHB-MIT dataset used, only 18 of the 23 patient sets. Exclusion criteria provided. |
| Bias in classification of interventions | Low | Multi-Scale-Rhythm Attention Feature Contrastive Learning well described. |
| Bias due to deviations from intended interventions | Low | Study carried out as intended. |
| Bias due to missing data | Low | No missing data reported. |
| Bias in measurement of outcomes | Low | Area under the curve (AUC), Sensitivity (Sn, the proportion of correct predictions to all seizure occurrences), False Prediction Rate (FPR/h, indicating the number of false alarms per hour), and p-value all reported on. |
| Bias in selection of the reported result | Low | All results reported. |
| **Overall Risk of Bias** | **Moderate** | Cohen's Kappa = 1.00 |

Table 12: *
Risk of Bias Assessment — Sarvi Zargar (2023)[12]

| Domain | Judgement | Justification |
|---|---|---|
| Bias due to confounding | Moderate | Age / sex reported but no covariate adjustment. |
| Bias in selection of participants | Low | 10 Epilepsia subjects used; inclusion criteria stated. |
| Bias in classification of interventions | Low | Deep transfer learning with MobileNetV2 + FC is described with preprocessing and input strategy. |
| Bias due to deviations from intended interventions | Low | Xception convolutional network with a Fully Connected (FC) classifier described. |
| Bias due to missing data | No Information | No information about excluded data provided. |
| Bias in measurement of outcomes | Low | Sensitivity and FPR reported; metrics computed for independent subjects. |
| Bias in selection of the reported result | Moderate | All results clearly displayed in Table 7. |
| **Overall Risk of Bias** | **Moderate** | Cohen's Kappa $= 0.73$ |

Table 13: *
Risk of Bias Assessment — Wang (2022)[13]

| Domain | Judgement | Justification |
|---|---|---|
| Bias due to confounding | Serious | No demographic adjustment. Only short pre-ictal and inter-ictal clips analysed. |
| Bias in selection of participants | Low | 12 patients were selected from CHB-MIT dataset who had less than 10 seizures per day consistent with current research and selection was justified. |
| Bias in classification of interventions | Low | Residual-convolution VAE architecture and training pipeline are fully described. |
| Bias due to deviations from intended interventions | Low | Procedure followed the published design; no deviations reported. |
| Bias due to missing data | No Information | No Information provided regarding missing data. |
| Bias in measurement of outcomes | Low | Accuracy, Sensitivity, FPR/h, AUC all used to measure outcomes. |
| Bias in selection of the reported result | Moderate | Only the best-case results are shown; no multiple-run statistics. |
| **Overall Risk of Bias** | **Serious** | Cohen's Kappa $= 0.42$ |

Table 14: *
Risk of Bias Assessment — Mustaqeem (2023)[14]

| Domain | Judgement | Justification |
|---|---|---|
| Bias due to confounding | Moderate | Paper claims a patient-independent "generalisation" model but does not analyse or adjust for inter-subject variability; no demographics or seizure-type controls. |
| Bias in selection of participants | Moderate | Only the first ten CHB-MIT subjects were used; no rationale for this subset. |
| Bias in classification of interventions | Low | Band-pass filtering and CNN architecture described in sufficient detail. |
| Bias due to deviations from intended interventions | Low | Training and testing followed the stated protocol with no reported deviation. |
| Bias due to missing data | No Information | Manuscript does not mention noisy or rejected EEG segments or any handling strategy. |
| Bias in measurement of outcomes | Low | Accuracy, sensitivity, and AUC are reported clearly. |
| Bias in selection of the reported result | Low | All the evaluation indicators are presented completely in Table 3, and there are no obvious selective reports. |
| **Overall Risk of Bias** | **Moderate** | Cohen's Kappa = 0.77 |

Table 15: *
Risk of Bias Assessment — Yu (2020)[15]

| Domain | Judgement | Justification |
|---|---|---|
| Bias due to confounding | Moderate | The study does not explicitly model or adjust for confounders, but dataset segmentation seems consistent. |
| Bias in selection of participants | Low | EEG from 21 patients used; preictal and interictal segments selected with consistent windows. |
| Bias in classification of interventions | Low | Local Mean Decomposition plus CNN pipeline described in sufficient detail. |
| Bias due to deviations from intended interventions | Low | Model follows planned pipeline. No deviation reported. |
| Bias due to missing data | No Information | EEG rejection and data cleaning procedures not described. |
| Bias in measurement of outcomes | Low | Sensitivity and FPR are reported; patient-independent protocol used. |
| Bias in selection of the reported result | Low | All the results are concentrated in Table 2 and Section 5, with no traces of selective reporting. |
| **Overall Risk of Bias** | **Moderate** | Cohen's Kappa = 0.53 |

Table 16: *
Risk of Bias Assessment — Duan (2019)[16]

| **Domain** | **Judgement** | **Justification** |
|---|---|---|
| Bias due to confounding | Serious | No confounder analysis performed; model tested without control for inter-patient variability or seizure types. |
| Bias in selection of participants | Moderate | Paper states CHB-MIT data but does not detail which subjects or how many recordings were included. |
| Bias in classification of interventions | Low | The model structure is detailed. Figure 1 shows the CRNN multi-scale module. |
| Bias due to deviations from intended interventions | Low | No indication of deviation; implementation aligned with described design. |
| Bias due to missing data | No Information | EEG quality control, missing segments, or artifact removal not discussed. |
| Bias in measurement of outcomes | Low | Acc, Sens, and Spec reported from a well-defined test set; appropriate seizure prediction metrics used. |
| Bias in selection of the reported result | Moderate | Only the main metric table is shown; auxiliary analyses are not provided. |
| **Overall Risk of Bias** | **Serious** | Cohen's Kappa = 0.78 |

Table 17: *
Risk of Bias Assessment — Nazari (2022)[17]

| **Domain** | **Judgement** | **Justification** |
|---|---|---|
| Bias due to confounding | Moderate | Model aims to improve few-shot seizure prediction, but does not explicitly control for inter-patient differences. |
| Bias in selection of participants | Low | Study transparently defines the target population (patients with rare seizures); patient-specific testing used. |
| Bias in classification of interventions | Low | CNN-based architecture and few-shot adaptation clearly described. |
| Bias due to deviations from intended interventions | Low | All experiments were carried out as designed and the process was not deviated from. |
| Bias due to missing data | Moderate | Unclear how sample was reduced to 15 from 18. Also states in abstract that the evaluation was from three patients. |
| Bias in measurement of outcomes | Low | Sensitivity and FPR were reported using realistic preictal detection; evaluation appears robust. |
| Bias in selection of the reported result | Low | The experimental results are presented truthfully and the data are listed in detail in Table 3. |
| **Overall Risk of Bias** | **Moderate** | Cohen's Kappa = 1.00 |

Table 18: *
Risk of Bias Assessment — Ntahobari (2022)[18]

| **Domain** | **Judgement** | **Justification** |
|---|---|---|
| Bias due to confounding | Moderate | No patient-specific information described aside from medication but no control stated. |
| Bias in selection of participants | Low | iEEG dataset clearly described; model applied across patient samples. |
| Bias in classification of interventions | Low | Extra Trees ensemble with SHAP optimisation well documented. |
| Bias due to deviations from intended interventions | Low | Model appears implemented as described. |
| Bias due to missing data | Low | The data integrity is good. Consecutive iEEG paragraphs are used and no significant missing data is reported. |
| Bias in measurement of outcomes | Moderate | Only AUC is reported; clinically critical metrics (e.g. sensitivity, false-alarm rate) are not provided. |
| Bias in selection of the reported result | Moderate | Reporting concentrates on speed-up and interpretability; ablations and variance measures are absent. |
| **Overall Risk of Bias** | **Moderate** | Cohen's Kappa = 0.75 |

Table 19: *
Risk of Bias Assessment — Zhao (2022)[19]

| **Domain** | **Judgement** | **Justification** |
|---|---|---|
| Bias due to confounding | Moderate | Combines human and canine iEEG without reporting age, medication, seizure-type or species-level adjustments; no stratified analysis, so confounders remain uncontrolled. |
| Bias in selection of participants | Low | Uses three public datasets (AES, Melbourne, CHB-MIT) exactly as released; no selective inclusion reported. |
| Bias in classification of interventions | Low | Neural architecture search and pruning strategy are clearly explained with compression pipeline diagrams. |
| Bias due to deviations from intended interventions | Low | No deviations reported; implementation strictly follows NAS-pruning setup. |
| Bias due to missing data | Low | Unreported data omissions or signal eliminations were not observed. The full-text experiments used complete records, and no obvious data shedding issues were observed. |
| Bias in measurement of outcomes | Low | Sensitivity and FAR reported; energy consumption measured to support wearable feasibility. |
| Bias in selection of the reported result | Low | All the main results were presented in Section IV and the charts, and no selective result reports occurred. |
| **Overall Risk of Bias** | **Moderate** | Cohen's Kappa = 0.26 |

Table 20: *
Risk of Bias Assessment — Xu (2020)[20]

| Domain | Judgement | Justification |
|---|---|---|
| Bias due to confounding | Moderate | Preictal/interictal separation used; no formal adjustment for demographic or clinical confounders. |
| Bias in selection of participants | Low | CHB-MIT and iEEG (Kaggle) datasets used; preprocessing pipeline is clearly reported. |
| Bias in classification of interventions | Low | Two-branch CNN structure is clearly described with kernel configurations and training setup. |
| Bias due to deviations from intended interventions | Low | No deviations documented; methodology aligns with architecture. |
| Bias due to missing data | Low | The dataset is complete and no missing cases have been reported. |
| Bias in measurement of outcomes | Low | Sensitivity and FAR reported using unseen test set; evaluation thresholding described. |
| Bias in selection of the reported result | Low | Clearly report key indicators such as sensitivity, FAR, and AUC. |
| **Overall Risk of Bias** | **Moderate** | Cohen's Kappa = 0.73 |

Table 21: *
Risk of Bias Assessment — Yang (2021)[21]

| Domain | Judgement | Justification |
|---|---|---|
| Bias due to confounding | Moderate | The model is evaluated across multiple patients but no demographic or seizure-type variables are analyzed or controlled. |
| Bias in selection of participants | Low | Public CHB-MIT dataset is used with consistent preprocessing and signal transformation. |
| Bias in classification of interventions | Low | Dual self-attention residual network is fully described, including residual blocks and attention modules. |
| Bias due to deviations from intended interventions | Low | No deviation reported; architecture and pipeline match description. |
| Bias due to missing data | Low | No obvious missing data was observed. Modeling was conducted using complete records. |
| Bias in measurement of outcomes | Low | Accuracy, sensitivity, specificity, and AUC reported using standard test data. |
| Bias in selection of the reported result | Moderate | Single configuration reported; cross-validation and hyperparameter robustness not evaluated. |
| **Overall Risk of Bias** | **Moderate** | Cohen's Kappa = 0.515 |

Table 22: *
Risk of Bias Assessment — Maddineni (2022)[22]

| **Domain** | **Judgement** | **Justification** |
|---|---|---|
| Bias due to confounding | Moderate | Some patient specific information provided but no controls stated. |
| Bias in selection of participants | Low | 21 patients from the CHB-MIT dataset used. |
| Bias in classification of interventions | Low | Hybrid transformer with FFT preprocessing is clearly described with architecture diagrams. |
| Bias due to deviations from intended interventions | Low | Implementation follows the proposed model setup. |
| Bias due to missing data | No Information | No explanation of EEG segment rejection, artifact handling, or quality control. |
| Bias in measurement of outcomes | Low | Sensitivity and FAR are reported; performance metrics are clinically relevant and on patient-level data. |
| Bias in selection of the reported result | Moderate | Results shown for a single configuration; uncertainty measures or alternative setups not discussed. |
| **Overall Risk of Bias** | **Serious** | Cohen's Kappa = 0.467 |

Table 23: *
Risk of Bias Assessment — Varnosfaderani (2024b)[23]

| **Domain** | **Judgement** | **Justification** |
|---|---|---|
| Bias due to confounding | Moderate | No demographic or treatment adjustment despite cross-validation. |
| Bias in selection of participants | Low | 18/23 CHB-MIT subjects used; exclusion criteria stated. |
| Bias in classification of interventions | Low | Multi-Scale-Rhythm Attention Feature Contrastive Learning well described. |
| Bias due to deviations from intended interventions | Low | Training/evaluation followed stated protocol. |
| Bias due to missing data | Low | Authors report no missing EEG after preprocessing. |
| Bias in measurement of outcomes | Low | AUC, Sensitivity, FPR/h, p-value reported on held-out folds. |
| Bias in selection of the reported result | Moderate | Only final pipeline reported; no ablation or alternative variants. |
| **Overall Risk of Bias** | **Moderate** | Cohen's Kappa = 0.515 |
